# *ACE2* and *TMPRSS2* variants and expression as candidates to sex and country differences in COVID-19 severity in Italy

**DOI:** 10.1101/2020.03.30.20047878

**Authors:** Rosanna Asselta, Elvezia Maria Paraboschi, Alberto Mantovani, Stefano Duga

**Affiliations:** Department of Biomedical Sciences, Humanitas University, Via Rita Levi Montalcini 4, 20090 Pieve Emanuele, Milan, Italy; Humanitas Clinical and Research Center, IRCCS, Via Manzoni 56, 20089 Rozzano, Milan, Italy; The William Harvey Research Institute, Queen Mary University of London, London EC1M 6BQ, UK

**Keywords:** SARS-CoV-2, COVID-19, *ACE2*, *TMPRSS2*, genetic variants, Italian population

## Abstract

**Background:** As the outbreak of coronavirus disease 2019 (COVID-19) progresses, prognostic markers for early identification of high-risk individuals are an urgent medical need. Italy has the highest rate of SARS-CoV-2 infection, the highest number of deaths, and the highest mortality rate among large countries. Worldwide, a more severe course of COVID-19 is associated with older age, comorbidities, and male sex. Hence, we searched for possible genetic components of the peculiar severity of COVID-19 among Italians, by looking at expression levels and variants in *ACE2* and *TMPRSS2* genes, which are crucial for viral infection.

**Methods:** Exome and SNP array data from a large Italian cohort representative of the country’s population were used to compare the burden of rare variants and the frequency of polymorphisms with European and East Asian populations. Moreover, we looked into gene expression databases to check for sex-unbalanced expression.

**Results:** While we found no significant evidence that *ACE2* is associated with disease severity/sex bias in the Italian population, *TMPRSS2* levels and genetic variants proved to be possible candidate disease modulators, contributing to the observed epidemiological data among Italian patients.

**Conclusions:** Our analysis suggests a role for *TMPRSS2* variants and expression levels in modulating COVID-19 severity, a hypothesis that fosters a rapid experimental validation on large cohorts of patients with different clinical manifestations.

## INTRODUCTION

As we write, Italy, Europe, and the entire world are facing one of the worst medical emergencies spanning centuries, the coronavirus disease 2019 (COVID-19) pandemia due to infection by SARS-CoV-2 virus. The early identification of risk factors for COVID-19 is an urgent medical need to provide the appropriate support to patients, including access to intensive care units.

Presently, Italy has one of the highest rate of SARS-CoV-2 infection in the world among large countries, with 143 cases per 100,000 people, the highest number of deaths and the highest mortality rate, 10.5% vs. an average value of 4.6% (as of March 28th, 2020, data from https://coronavirus.jhu.edu/map.html). These data may have different explanations, including: 1) the number of tests performed, 2) the structure of the population (Italy has the oldest population in Europe) [https://ec.europa.eu/eurostat/data/database], 3) the percentage of smokers, even though no significant association was found between smoking and severity of COVID-19 in a very recent study on the Chinese population,^1^ 4) the possible existence of a different virus strain,^2^ 5) the concentration of severe cases in a limited region of the country, potentially overwhelming the available intensive care units, as well as 6) differences in environmental factors (e.g. air pollution). However, there could also be some peculiar genetic characteristics of the Italian population that may have an impact on the susceptibility to viral infection, the disease severity, and the number of patients shedding huge amounts of virus.

What is unquestionable is a more severe course of the disease associated with older age and high number of comorbidities and with the male sex (male:female ratio in case fatality rate among Italians 1.75, data from the Italian National Institute of Health: https://www.epicentro.iss.it/coronavirus/), a feature shared with the 2003 SARS epidemic and MERS.^3-5^ Indeed, while men and women have similar susceptibility to both SARS-CoV-2 and SARS-CoV, men are more prone to have higher severity and mortality, independently of age.^3^ Among the many possible factors impacting on sex-related differences in disease manifestations, including the fact that females are known to mount a stronger immune response to viral infections compared to males due to more robust humoral and cellular immune responses,^6^ we decided to center our attention on possible genetic components, with a particular focus on the Italian population.

It was recently demonstrated that both angiotensin I converting enzyme 2 (ACE2) and the transmembrane protease, serine 2 (TMPRSS2) are crucial for SARS-CoV-2 entry into host cells.^7,8^ While ACE2 is the main receptor for the spike (S) protein of both SARS-CoV and SARS-CoV-2, mediating viral attachment to target cells, TMPRSS2 cleaves protein S at the S1/S2 and the S2 ‘sites, allowing fusion of viral and cellular membranes.^8^ Both genes have been proposed to modulate susceptibility to SARS-CoV,^9,10^ and are good candidates to mediate sex-related effects: *ACE2* is located on the X chromosome, while *TMPRSS2* expression is responsive to androgen/estrogen stimulation.^11^

## METHODS

### Gene expression data

Expression data for *ACE* and *TMPRSS2* genes were obtained through the: 1) genotype-tissue expression (GTEx) database (https://gtexportal.org/home/), which was also used to extract quantitative trait loci (eQTLs) for the two genes (all data based on RNAseq experiments); and 2) Gene Expression Omnibus (GEO) repository (https://www.ncbi.nlm.nih.gov/geo/). In particular, two GEO datasets were extracted and analyzed: 1) GSE66499, reporting microarray data on 152 normal lung samples from Caucasian individuals; 2) GSE19804, reporting microarray data on 60 normal lung samples from Taiwanese females.

### Genetic data

Genetic data for general European and East Asian populations were retrieved through the GnomAD repository, which contains data on a total of 125,748 exomes and 71,702 genomes (https://gnomad.broadinstitute.org/). As for Italians, details on whole-exome sequencing (on 3,984 individuals) and genome-wide microarray genotyping (on 3,284 individuals) of the analyzed cohort are specified elsewhere.^12-14^ Imputation procedures are detailed in Supplementary materials.

### Statistical analysis

Expression levels were compared by using either the Kruskal-Wallis test (RNAseq data) or the student t test (microarray data). Allele frequencies were compared using the chi square test. All calculations were performed using the R software (https://www.r-project.org/). P values are presented as non-corrected for multiple testing.

## RESULTS AND DISCUSSION

### ACE2

For most X-chromosome genes, the double allelic dosage in women is balanced by the epigenetic silencing of one of the X chromosomes in early development.^15^ However, the X-chromosome inactivation (XCI) is incomplete in humans and up to one third of genes are expressed from both alleles, with the degree of XCI escape varying between genes and individuals.^16^ *ACE2* is one of the genes escaping X inactivation, but it belongs to a subgroup of X-chromosome genes escaping XCI showing an uncharacteristically heterogeneous pattern of male-female expression, with higher expression in men in several tissues.^17^ Specifically concerning the lung, a recent analysis on published expression data, reported a substantial similar level of *ACE2* transcript in men and women,^18^ however, another study, using single-cell sequencing, found a higher expression of *ACE2* in Asian men.^19^ Supplementary Figure 1 reports data on *ACE2* mRNA expression levels in the lung as retrieved from the largest datasets available in the literature; no substantial differences were found between males and females, nor between younger and older women, thus confirming what already observed by Cai and colleagues.^18^

**Figure 1:**
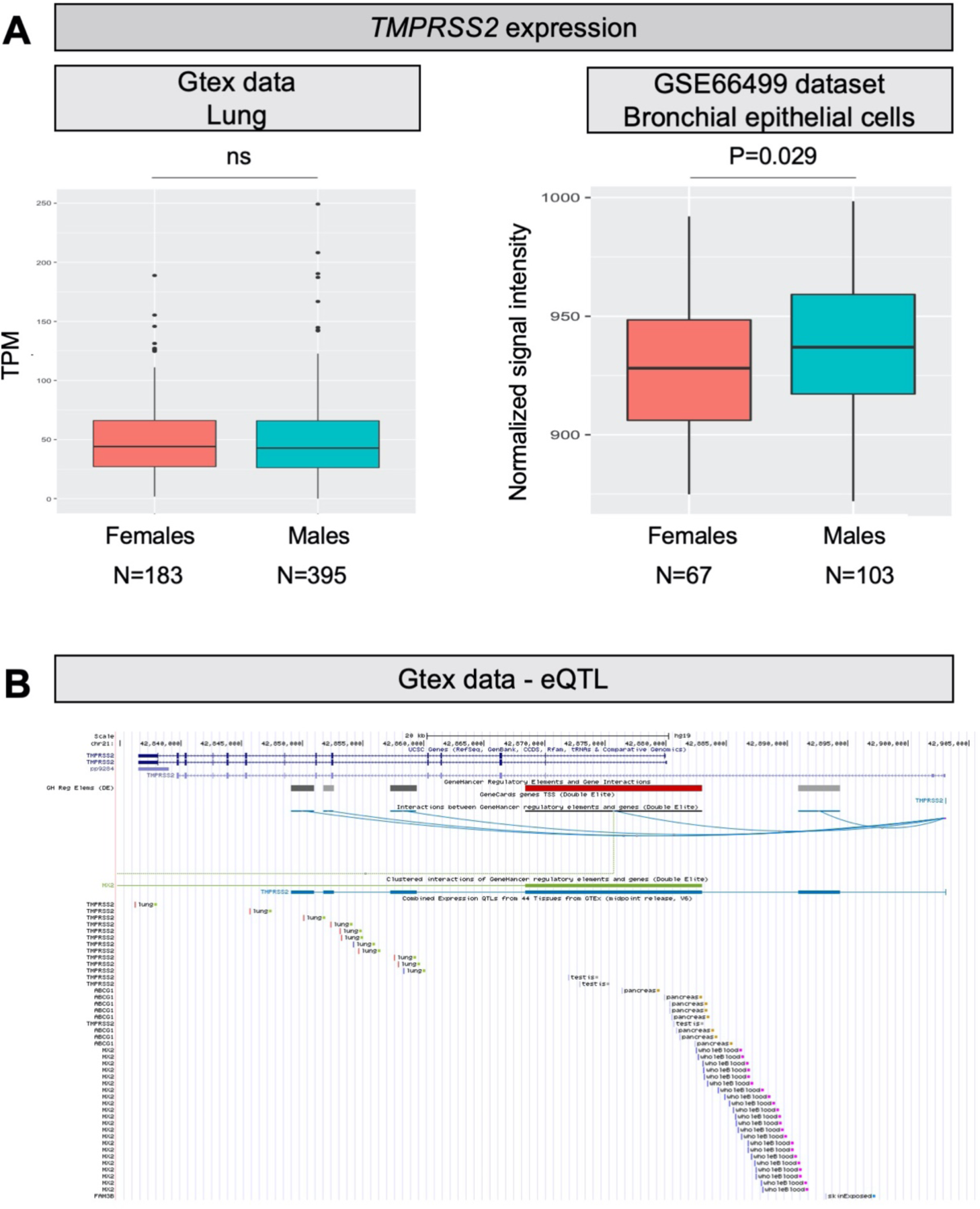
*TMPRSS2* expression levels and eQTLs. **A)** Both panels show *TMPRSS2* mRNA expression levels in human normal lung samples stratified according to sex. On the left, data were retrieved for a total of 578 RNAseq experiments from the GTex repository. Expression levels are reported as transcripts per kilobase million (TPM). On the right, data were collected for a total of 170 microarray experiments from the GEO database. Expression levels are reported as normalized signal intensities. P values were calculated by using either the Kruskal-Wallis or the student t test. **B)** Screenshot from the UCSC Genome browser (http://genome.ucsc.edu/; GRCh37/hg19) highlighting the *TMPRSS2* region (coordinates chr21: 42,835,000-42,905,000). The panel shows the following tracks: i) the ruler with the scale at the genomic level; ii) chromosome 21 nucleotide numbering; iii) the UCSC RefSeq track; iv) enhancers (grey and red bars) from GeneHancer database; v) interactions (curved lines) connecting GeneHancer regulatory elements and genes: all curved lines converge towards the androgen-responsive enhancer for the *TMPRSS2* gene described by Clinckemalie and colleagues.^28^

Another possible sex-related effect might be due to the fact that males are hemizygous for the gene, therefore, in the presence of an *ACE2* allelic variant increasing disease susceptibility or severity, men will have all cells expressing the risk variant. Based on this hypothesis, we looked into the genetic variation in *ACE2*. A recent manuscript explored this same topic in different populations using data from public databases. ^20^ However, a specific analysis of the Italian population is Iacking.

We have therefore exploited the available data on 3,984 exomes obtained from an Italian cohort representative of the whole country^12,13^ to extract the variants in exons and splice junctions of *ACE2*. Variants were filtered for quality and classified according to their predicted effect at protein level and on splicing. Concerning rare variants (i.e. those with a minor allele frequency, MAF, <1%; to be used in burden tests), we considered only null variants, abolishing or significantly impairing protein production (nonsense, out-of-frame ins/dels, and splicing variants), and missense variants predicted to be deleterious or possibly deleterious by all the 5 prediction algorithms used (see Supplementary Methods). Concerning common variants (i.e., MAF>5%), all were retained for comparing their frequency with those of the European (non Finnish) and East Asian populations, retrieved from the GnomAD repository.

No significant differences in the burden of rare deleterious variants were observed comparing the Italian population with Europeans and East Asians (Table 1A). Concerning common exonic variants, the only striking difference, as also noticed by Cao and colleagues,^20^ was observed for the single nucleotide polymorphism (SNP) rs2285666 (also called G8790A), with the frequency of the rare A allele being 0.2 in Italians and Europeans, and 0.55 in East Asians (P=2.2*10-16 for difference in Italians vs East Asians; Table 1B). This variant was extensively studied as a potential risk factor for hypertension, type 2 diabetes, and coronary artery disease,^21,22^ hence possibly constituting a predisposing factor also for the comorbidities observed in COVID-19 patients. A single paper reports the association of the three rs2285666 genotypes with ACE2 protein level measured in serum by ELISA, with the A/A genotype having an expression level almost 50% higher than the G/G genotype.^23^ Given the position of the variant, at nucleotide +4 in the donor splice site of intron 3 (c.439+4G>A), we calculated the predicted effect on splicing and indeed the substitution of G with an A is predicted to increase the strength of the splice site of about 9.2% (calculation made through the Human Splicing Finder v.3.1 webtool, http://www.umd.be/HSF/), consistently with the higher level of ACE2 protein in serum. It would be crucial to compare the frequency of this variant with *ACE2* expression in the lung and with susceptibility to viral infection and severity of COVID-19 manifestations. Of note, no eQTL for *ACE2* in the lung has been described so far in the GTEx database, and investigations on this topic are recommended.

**Table 1A.** Burden of rare mutations in the *ACE2* gene in different populations.

| Population | N alleles | T1 | Freq T1 | ITA | EUR | EAS |
| --- | --- | --- | --- | --- | --- | --- |
| ITA | 4422 | 7 | 0.0016 | - | P=0.518 | P=0.974 |
| EUR | 92545 | 200 | 0.0022 | P=0.518 | - | P=0.077 |
| EAS | 14840 | 21 | 0.0014 | P=0.974 | P=0.077 | - |

Total allele counts, carrier allele counts, and carrier frequencies are shown; only deleterious variants with MAF less than 1% were considered in the burden analysis. The ‘deleterious’ set is defined by missense variations predicted to be possibly damaging by all the 5 algorithms used (LRT score, MutationTaster, PolyPhen-2 HumDiv, PolyPhen-2 HumVar, and SIFT), and loss-of-function variants (nonsense, frameshift, and splicing variants affecting the donor/acceptor sites).
T1: alleles carrying damaging variants; Freq T1: frequency of T1 allele; ITA: Italian population; EUR: European population; EAS: East Asian population

**Table 1B.** Common exon variants in the *ACE2* gene in different populations.

| Variant ID | Consequence | A1/<br>N alleles<br>ITA | Freq<br>ITA | A1/<br>N alleles<br>EUR | Freq<br>EUR | A1/<br>N alleles<br>EAS | Freq<br>EAS | ITA<br>Vs<br>EUR | ITA<br>Vs<br>EAS | EUR<br>Vs<br>EAS |
| --- | --- | --- | --- | --- | --- | --- | --- | --- | --- | --- |
| rs2285666 | c.439+4G>A | 909/<br>4408 | 0.206 | 17240/<br>86164 | 0.200 | 7336/<br>13387 | 0.548 | 0.331 | P< 2.2e-16 | P< 2.2e-16 |
| rs35803318 | p.Val749Val | 235/<br>4422 | 0.053 | 3935/<br>88946 | 0.044 | 0/13918 | 0.0 | P=0.0058 | P< 2.2e-16 | P< 2.2e-16 |

Total allele counts, carrier allele counts, and carrier frequencies are shown; only variants with MAF more than 5% were considered.
A1: alleles carrying variants; Freq A1: frequency of A1 allele; ITA: Italian population; EUR: European population; EAS: East Asian population

### TMPRSS2

Our second candidate is *TMPRSS2*. This gene is well known to oncologists as genetic rearrangements producing a fusion between *TMPRSS2* and *ERG* (or, more rarely, other members of the ETS family) are the most frequent genetic lesions in prostate cancer patients.^24^ As *TMPRSS2* is an androgen responsive gene, the fusion results in androgen dependent transcription of ERG in prostate tumor cells. Therefore, we can hypothesize that men might have higher *TMPRSS2* expression also in the lung, which might improve the ability of SARS-CoV-2 to enter cells by promoting membrane fusion. Looking into GTEx and GEO data, the overall expression of *TMPRSS2* in the lung is only slightly increased in men (P=0.029; Figure 1A). However, *TMPRSS2* expression is also promoted by estrogens,^11^ and therefore the situation might be different when considering individuals above 60 years, who are at higher risk of fatal events due to COVID-19, as in this group women will all be postmenopausal. According to this hypothesis, we checked the expression of the gene in lungs of men and females at different ages, but no substantial differences emerged between males and females (neither below, nor above 60 years of age; data not shown).

Finally, we explored genetic variation in *TMPRSS2* in search of variants, possibly already annotated as eQTL in the lung, which might have an impact on the serine protease expression as well as on its catalytic activity. Again, we used the available Italian exome data, as well as data deposited in GnomAD.^25^

Firstly, we looked at the overall burden of deleterious rare variants, using the variant classification described above. Italians had a significant decrease in the burden of deleterious variants compared to Europeans (P=0.039; Table 2A) suggesting that they might have a higher level of TMPRSS2 protein/activity, which in principle should be a risk factor for more severe disease course. This is even more true for the East Asian population, however, in this case, we must consider that the number of individuals over 65 years of age in Italy is more than double the one in the Hubei province (22.7 vs. 10%, respectively) and this is a major determinant of disease lethality.

**Table 2A.**
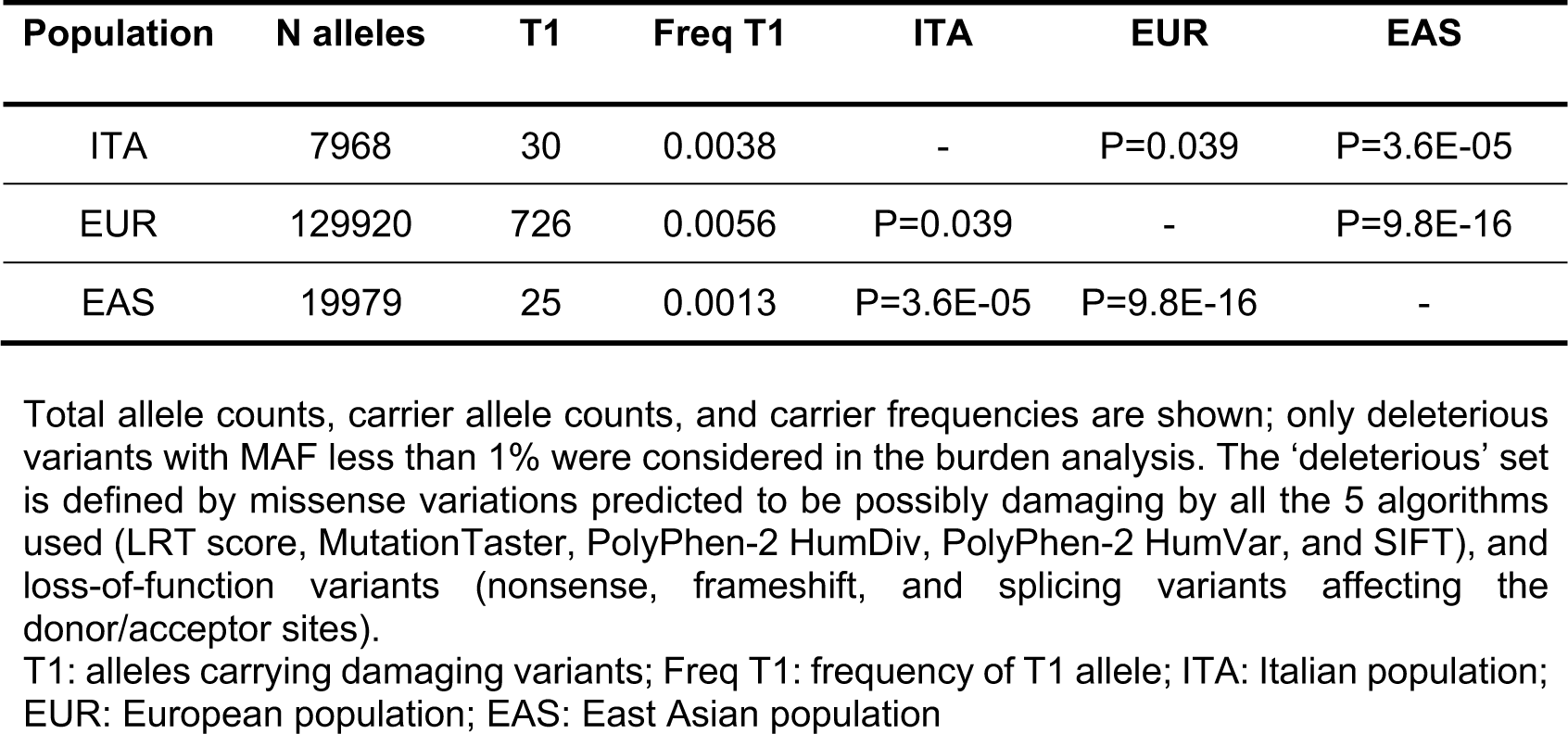
Burden of rare mutations in the *TMPRSS2* gene in different populations.

Focusing specifically on common exonic variants, 4 SNPs showed significantly (P<2.2*10-16) different frequencies when comparing the Italian population with East Asians (and with Europeans) (Table 2B); 3 of them are synonymous variants, whereas one is the missense substitution p.Val160Met, which impacts on a residue far from the serine protease catalytic triad. This variant was previously found significantly associated with genomic rearrangements involving *TMPRSS2*, with the risk of prostate cancer^26^ and with shorter time to prostate cancer diagnosis for high-risk patients.^27^ Concerning eQTLs, a number of variants significantly impacting on *TMPRSS2* expression in the lung (GTex data) are reported in the 3’ region of the gene (Figure 1B). In Table 2C, a list of the most significant (P<1*10-8), together with their GnomAD frequencies in the East Asian and European populations, are reported. As for the Italian frequencies, we took advantage of the genome-wide association study (GWAS) performed on the above-described cohort (for a total of 3,284 individuals);^14^ in this case, we had to infer genotype frequencies by an imputation approach (for details, see Supplementary Methods). Interestingly, all these eQTLs appear to have extremely different frequencies among populations. In particular, 2 different haplotypes can be inferred from frequency data:

**Table 2B.**
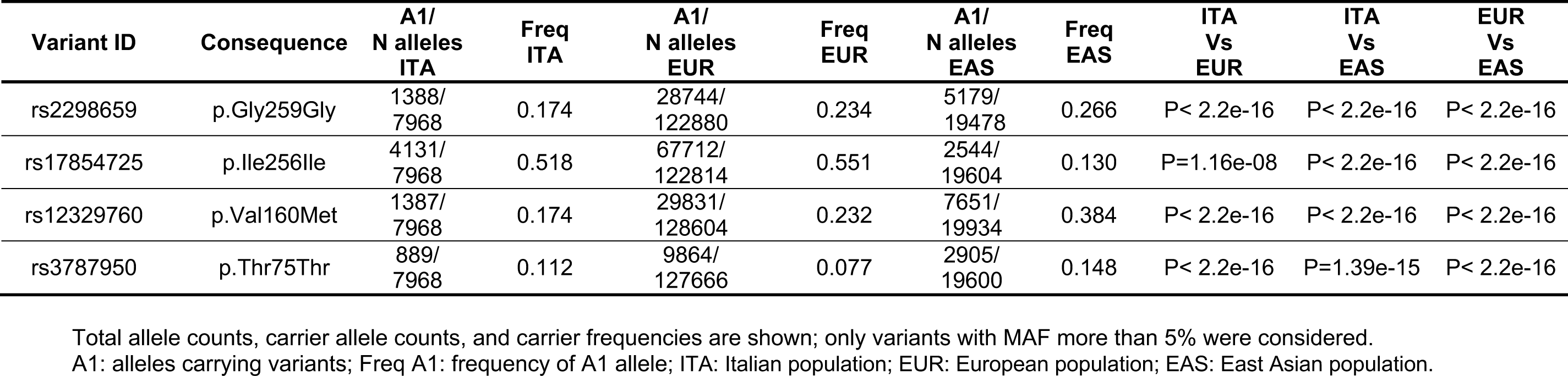
Common exon variants in the *TMPRSS2* gene in different populations.

**Table 2C.** eQTL variants in the *TMPRSS2* gene in different populations.

| Variant ID | P<br>GTEx | NES<br>GTEx | Freq<br>ITA | Freq<br>EUR | Freq<br>EAS | ITA vs<br>EUR | ITA vs<br>EAS | EUR vs<br>EAS |
| --- | --- | --- | --- | --- | --- | --- | --- | --- |
| rs463727 | 5.0e-10 | 0.12 | 0.44 | 0.46 | 0.0051 | P=0.038 | P<2.2e-16 | P<2.2e-16 |
| rs2070788 | 8.9e-9 | -0.11 | 0.55 | 0.53 | 0.66 | P=0.003 | P=4.7e-15 | P<2.2e-16 |
| rs9974589 | 7.4e-9 | -0.12 | 0.55 | 0.53 | 0.66 | P=0.002 | P=3.3e-15 | P<2.2e-16 |
| rs34624090 | 9.2e-9 | 0.12 | 0.43 | 0.45 | 0.0051 | P=0.005 | P<2.2e-16 | P<2.2e-16 |
| rs7364083 | 3.3e-9 | -0.12 | 0.56 | 0.53 | 0.65 | P=8.7e-05 | P=1.9e-10 | P<2.2e-16 |
| rs55964536 | 1.9e-9 | 0.12 | 0.46 | 0.49 | 0.0045 | P=4.6e-04 | P<2.2e-16 | P<2.2e-16 |
| rs734056 | 1.3e-9 | 0.12 | 0.47 | 0.49 | 0.0051 | P=0.030 | P<2.2e-16 | P<2.2e-16 |
| rs4290734 | 8.3e-10 | 0.12 | 0.47 | 0.49 | 0.0051 | P=0.019 | P<2.2e-16 | P<2.2e-16 |
| rs34783969 | 3.9e-10 | 0.12 | 0.47 | 0.49 | 0.0051 | P=0.027 | P<2.2e-16 | P<2.2e-16 |
| rs11702475 | 8.4e-10 | 0.12 | 0.47 | 0.49 | 0.0046 | P=0.015 | P<2.2e-16 | P<2.2e-16 |
| rs35899679 | 7.8e-9 | 0.11 | 0.44 | 0.46 | 0.0051 | P=0.004 | P<2.2e-16 | P<2.2e-16 |
| rs35041537 | 3.6e-9 | 0.12 | 0.44 | 0.47 | 0.0051 | P=8.6e-04 | P<2.2e-16 | P<2.2e-16 |

NES: normalized effect size; Freq: frequency of the minor allele; ITA: Italian population; EUR: European population; EAS: East Asian population

1. A frequent “European” haplotype (composed at least of SNPs rs463727, rs34624090, rs55964536, rs734056, rs4290734, rs34783969, rs11702475, rs35899679, and rs35041537), which is totally absent in the Asian population. Interestingly, this haplotype has been functionally linked to another eQTL (rs8134378), located at a known androgen-responsive enhancer for *TMPRSS2*, 13 kb upstream of the *TMPRSS2* transcription start site^28^ (Figure 1B). Hence, this haplotype is expected to up regulate *TMPRSS2* gene expression in an androgen-specific way.
2. A second haplotype, predicted to be associated with higher *TMPRSS2* expression, is characterized by 3 SNPs (rs2070788, rs9974589, rs7364083), whose MAF is significantly increased in Europeans (9% increase in Italians respect to East Asians, P<1.9*10^−10^). Importantly, a small-scale GWAS, comparing the distribution of genetic variants in severe and mild cases of patients with A(H1N1)pdm09 influenza, identified rs2070788 as being associated with increased risk to both human A(H7N9) and severe A(H1N1)pdm09 influenza.^10^ Of note, also in A(H7N9) influenza, the proportion of male patients was more than double that of female patients.^29^

## Limitations and conclusions

We are aware of the limitations of our study: first of all we focused our attention only to two candidate genes identified on the basis of their crucial role in viral infection and on the a priori probability that they might mediate sex-specific effects. A number of other X-linked genes (such as *IL13, IL4, IL10, XIST, TLR7, FOXP3)* and Y-linked genes (*SRY, SOX9*) may underlie sexually dimorphic immune responses.^30^ Moreover, the number of non-genetic determinants of sex-biased severity and case fatality rates is huge and probably has to do not only with sex differences in both innate and adaptive immune responses,^6^ but also with gender and cultural habits in different countries.

In conclusion, we have explored possible genetic components impacting on the sex-biased severity of COVID-19, focusing on effects mediated by *ACE2* and *TMPRSS2* genes in the Italian population. From available data, it seems unlikely that sex-differences in *ACE2* levels can explain sex differences in disease severity. However, it remains to be evaluated if changes in *ACE2* levels in the lung correlate with susceptibility and severity of SARS-CoV-2 infection. Experimental data from patients with different disease manifestations are urgently needed. Among the analyzed hypotheses, the most interesting signals refer to sex-related differences in *TMPRSS2* expression and in genetic variation in *TMPRSS2*. In particular, we identified an exonic variant (p.Val160Met) and 2 distinct haplotypes showing profound frequency differences between East Asians and Italians. The rare alleles of these haplotypes, all predicted to induce higher levels of *TMPRSS2*, are more frequent in the Italian than in the East Asian population; in one case, the haplotype could be regulated through androgens (possibly explaining the sex bias in COVID-19 severity?), in the other case, a SNP belonging to the haplotype has been associated with increased susceptibility to influenza, possibly related to a higher susceptibility in Italians and Europeans. Our data, beside suggesting possible explanations for the unusually high, relative to known data, lethality rates among Italians, provide reference frequencies in the general Italian population for candidate variants that can be compared to genetic data from patients infected by SARS-CoV-2 with different disease manifestations, as soon as they will be available on large numbers of patients. These studies will hopefully help to identify useful prognostic markers to stratify patients and provide the best care to high-risk individuals.

## Data Availability

Data used for the analysis are already available on research databases

## FUNDING

This work was supported by Ricerca Corrente (Italian Ministry of Health), intramural funding (Fondazione Humanitas per la Ricerca). A generous contribution of the Dolce&Gabbana Fashion Firm is gratefully acknowledged.

## AUTHORS’ CONTRIBUTIONS

All authors contributed to the study design. EMP did the genetic analysis, RA performed the statistical analysis, SD drafted the manuscript and supervised the entire study. All authors critically reviewed the manuscript and approved the final draft.

## CONFLICT OF INTEREST STATEMENTS

No conflict of interest to disclose.

## Notes

### Competing Interest Statement

The authors have declared no competing interest.

